# The mediating role of social network between social trust and life satisfaction of older people: A structural equation model analysis approach

**DOI:** 10.1101/2024.06.09.24308669

**Authors:** Xinyu He, Zhaoyong Yang, Haimei Li, Shihe Li, Chuanxin Xu

**Author notes:** **Correspondence:** Xinyu He; Haimei Li.

## Abstract

**Background:** Although studies have demonstrated the relationship between the social trust and life satisfaction of older people, but existing studies have rarely considered the fewer studies have investigated social network as a mediating variable between social trust and life satisfaction.

**Methods:** In this study, 297 participants (age: ≥60 years) in Chengdu, Sichuan Province, China, completed a questionnaire of social trust, social network, and life satisfaction. Structural equation model analysis and bootstrap methods were used to prove the mediating effect of social network.

**Results:** Social trust was positively related to life satisfaction (*B* =0.410; p <0.01) and social network (*B* =0.472; p <0.01), social network was positively related to life satisfaction (*B* =0.433; p <0.01). and social network played an intermediary role in the relationship between social trust and life satisfaction (indirect effects = 0.321, bootstrap 95% CI = [0.051, 0.968]).

**Conclusion:** These findings suggest the importance of social trust for life satisfaction, with social network acting as a positive mediator, particularly in China. In conclusion, this study exploring the mediating effect of social network provides knowledge essential for ensuring a happy old age. The study results can help advance psychological research on life satisfaction of older people and suggest new intervention strategies in this field. We encourage increasing the expansion of social networks through various means to enhance the impact of social trust on life satisfaction of older people.

## Background

According to China’s seventh census, the number of people aged ≥ 60 years has reached 264 million, accounting for 18.3% of the total population in 2020.^1^ By 2050, Chinese older people are estimated to account for one-third of the country’s population [1]. With the increase in the aging population, the question is whether they can have a good quality of life and whether a more active social interaction (measured as social network) can translate into higher life satisfaction levels [2, 3]. Life satisfaction, a useful concept in the study of social psychology well-being which reflects the cognitive aspect of subjective well-being, is associated with physical and mental health of older adults [4], which refers to one’s self-evaluation of his/her life [5, 6]. For example, a self-evaluation in the past, present, and future [7–9]. Consequently, life satisfaction of the elderly people has recently received considerable attention [10]. Understanding factors that impact life satisfaction not only can help understanding happiness and quality of life but also have important implications, as higher life satisfaction is related to better physical and mental health outcomes, such as a reduced risk of disease, depression and loneliness [11–13].

To study the factors of life satisfaction, several theoretical models have been developed, including a bottom-up, top-down and comprehensive accounts, which means that life satisfaction is made up of demographics (e. g., health, income, job), satisfaction with aspects of life (e. g., satisfaction with family or friendship, social life), and personality traits [14, 15]. Nonetheless, life satisfaction must be understood in the context of each culture [15]. In the collectivist culture, such as in China, people lay more emphasis on social relationships and connectedness with others [16], and social relationships are related to life satisfaction of the older people [17, 18]. However, with age and retirement, people more likely lose original social roles, and according to social disengagement theory, older people rarely interact with society and are willing to withdraw from their social system [19, 20]. To take new social roles and maintain positive social interactions, building new social networks and relationships are important to older people [21]. Knowing the concept of social capital, and more broadly, the significance of social relationships as a cornerstone of a functional society is crucial, because they have significant positive effects on life satisfaction [22, 23].

Social capital, which can be viewed from an individual’s perspective and considered as crucial social resource embedded in a network of community social relations, through which people share memberships, values, and social norms [24]. We considered individual-level social capital as a multi-dimensional concept, that can be measured on the basis of structural and cognitive dimensions [25]. Structural social capital, mainly refers to the objective social structure, such as social network and social participation. While cognitive social capital, refers to trust, values, beliefs, reciprocity. and other psychological processes [26, 27]. Some studies have proven that a close relationship exists between structural, cognitive social capital, and life satisfaction among older people [28, 29]. For example, according to a survey, social network has positive effects on life satisfaction [30]. More specifically, older people connect with each other through social networking, social trust and establishment of social relationships, which is helpful for improving life satisfaction [31, 32].

Although these associations are well known, few studies have explored the relationship between structural social capital and cognitive social capital, and, in particular, the path from social trust to life satisfaction through social network in the elderly people has not been fully explored. To fill the research gap, this study explored the mediating role of social network between the social trust and life satisfaction of older people.

### Social trust and life satisfaction

Social trust is a major indicator of cognitive social capital, which has been considered a key determinant of happiness [33–37]. Studies have recognized two types of social trust: interpersonal trust and institutional trust [38]. Interpersonal trust refers to individual’s psychological state of believing that most people can be trusted, including positive expectations about others’ behaviors[39, 40]. Trust facilitates a safe and stable social environment in relationships by reducing feelings of worry or anxiety [41]. Using multiple linear regression and a mixed-design analysis of variance, a survey tested a sample of community dwelling participants (age: 55-89 years) in the New Zealand. The results showed that the neighborhood relationship was associated with older people’s subjective well-being [42]. The more you trust your neighbors, the higher your life satisfaction will be. Using a sample of Chinese older people from the rural area, a study research showed that interpersonal trust and family support were significantly positively associated with life satisfaction [28].

Institutional trust reflects people’s attitudes and satisfaction toward state institutions (e.g., government institutions, educational institutions, medical institutions and judicial branches). Compared with young people, the trustworthiness of public institutions affects elderly people’s evaluation of society and thus their well-being [43]. On measuring the levels of trust of participants in eight institutions, a researcher found that institutional trust was positively associated with life satisfaction [17], another researcher investigated the predictive effect of generalized trust on health, happiness, and life satisfaction among older adults who lived in six non-Western countries, they reported that generalized trust can predict greater health and life satisfaction [44]. Exploring cross-sectional data of 5000 older people (≥ 65 years), the results of a study showed that community trust can mitigate inequalities in well-being and maintain good mentality [45]. Older people, with higher levels of institutional trust tend to exhibit better adherence and acceptance of policy measures and better satisfaction with their life [46]. According to the above literature, we propose the following hypothesis.

**Hypothesis 1** Social trust is positively related to life satisfaction in older people.

### Social network and life satisfaction

Social network is a crucial indicator of structural social capital, which is-“the array of social connections that give access to instrumental and emotional supports” [47]. The results of a study exhibited that social network size and network contact frequency could allow older people access beneficial resources and support, thereby helping them sustain higher life satisfaction [48]. Meanwhile, formal and informal social networks are the base from which social capital is generated to influence people’s behaviors and values, including well-being [49, 50]. On the source of happiness, the researchers found that a strong informal social network was associated with individual’s happiness, including how many times they see friends and relatives [12, 51]. This is because, older people who had a wide range of social relationships, were the happiest [52]. But another study indicated that the number of close friends had the weakest relationship with life satisfaction among older people [6]. Moreover, older people participating in social activities can expand their formal social network, and thus improve their life satisfaction [53–55]. The findings of researcher suggested that group participation through formal social networks is linked to life satisfaction in later life [56]. Similarly, another researcher analyzed the association between social network and quality of life in older Chilean people and found that participating in formal social support networks is associated with high quality of life [57].

Chinese cultural values may also be a crucial player in explaining how social network can affect life satisfaction of older people. East Asian cultures tend to be more collectivist and communal [58, 59]. Collectivism, a unique cultural context in Chinese society, values connections with others and social relationships. In this context, social relationships, such as family and friend-focused network types, highly affect Chinese people’s life satisfaction, especially that of older people, who are deeply influenced by the Chinese traditional culture [60, 61].

According to the socioemotional selectivity theory [62], those who grow older tend to choose social networks that allow them to form close social relationships. Thus, compared with young people, older people are more likely to choose social networks on which they can trust and that provide opportunities for companionship. On the one hand, older people are embedded in “family-centered” and “friend-centered” social networks from where they receive social support [63]. The more frequent the face-to-face contact with other network members, the more social support people can obtain [48]. On the other hand, participating in formal social networks and meaningful community activities can bring a sense of belonging and self-identity, as well as provide opportunities for social engagement and sharing leisure and entertainment activities. This, thus, enhances life satisfaction among older people [64]. The following hypothesis can be established.

**Hypothesis 2** Social network is positively related to life satisfaction in older people.

### Social trust and social network

Social trust is linked to social network. Using data from the first Norwegian survey, the study demonstrated that in non-kin relationships, social networks were associated with higher levels of generalized trust [65]. Another related feature of networks and trust relationships is possibly the resources accessible to individuals through their interactions [66, 67]. Studies have found that the higher the frequency of contact of the elderly people with the outside world, the higher is the number of resources they can access. Individual’s resources (family relationship, and network resources) and social trust are positively correlated [68–69, 30]. A research’s findings showed that if the level of social trust is higher, an individual would more likely approach the social network actively, thereby gaining more material and emotional support [70]. According to the above literature, we posit the following hypothesis.

**Hypothesis 3** Social trust is positively related to social network in older people.

### Mediation effect of social network on the relations between social trust and life satisfaction

The social capital theory can explain how social network can mediate the relationship between social trust and life satisfaction. Social capital can be considered to stem from a social network, which provides resources such as social support, trust, and information. This network, different from the social cohesion concept of social capital, can be evaluated at the individual or collective level [71]. Broadly speaking, approaches used for studying social capital suggest that social capital can be measured by the level of collective or individual resources [8]. Some researches focused on the study of social capital as a level of collective resources, while Lin and colleagues mainly focused on the impact of social capital as a level individual resource [72, 73]. Considering that the goal of the present study is to explore the mediating effect of the social network on the relationship between social trust and life satisfaction of older people, we applied Lin’s definition of social capital, that is, the sum of individual-level resources [49] (e.g., social support and, information). Also, social capital is the social network among individuals that is based on trust [70]. It refers to the ability of individuals to access resources by being a member of a particular social network, that is, to extract value from social relationships that can be mobilized. In other words, social capital is a resource embedded in a social network [74]. The influence of powerful and ties in a network is related to social capital. These social networks and ties can be of numerous types like “functional support, emotional support, a sense of belonging, or shared values [72, 75]. People communicate and connect with each other through social networking and establishing social relationships which can improve life satisfaction [59, 60]. It should be noted that social network is based on social trust, a higher level of life satisfaction is reported by people who believed and trusted other people for social supports in need of help [76]. In this sense, social trust may affect life satisfaction through social networks.

As people age, they experience a decline in cognitive and physical abilities, as well as a decrease in social contact [77, 78]. In addition, their dependency on others increase [79], and they may be more exposed to social isolation and thus are likely to be lonely and depressed [80, 81]. Therefore, they require additional social supports, and social network is the carrier of gaining that social support, more stable instrumental and affective supports can be gained through social networks [82]. Social networks are established through human interactions or communications (e.g., social participation) to realize the exchange of information (e.g., health information). They provide immediate emotional benefits to older people and enhance their sense of social support [83], thereby shaping their identity and self-worth [84]. Trust facilitates the establishment of social networks [70], older people have different relational networks, including family, relatives, friends, and a broader community. All of these networks are important for older people, because they provide opportunities for maintaining good interpersonal relationships and offer a sense of belonging, and feeling of being needed, and are conducive to improving their life satisfaction [85–87]. Notably, all good relationships are built on trust [88, 89]. More specifically, social capital of older people, which is based on social trust, and the expansion of social networks to accumulate social resources and social support, thus enhancing older people’s life satisfaction. Therefore, we hypothesize the following:

**Hypothesis 4** Social network plays a mediating role in the relationship between social trust and life satisfaction in older people.

## Methods

### Data

This study was conducted between December 20, 2016, and January 25, 2017 in five districts in Chengdu, namely: Jinjiang, Wuhou, Qingyang, Chenghua, and Jinniu. The study included people from 15 communities, as participants (age: ≥60 years). The name of the community was obtained from the official website of the Chengdu government, and the communities in each administrative region were numbered in an alphabetical order, The communities numbered 01, 11, and 21 in each administrative region were selected. Then a random sample of 20 people in each community and 60 people in each administrative region were surveyed. In total, 300 people in five administrative regions were selected for the questionnaire survey, and 297 valid questionnaires were collected. Finally, 297 Chinese people with help and explanation as required from professional researchers, completed the survey that formed the basis of this study. The survey included questions on socio-demographic information, in addition to the variables social trust, social network, and life satisfaction. This survey was completely anonymous and approved by the Ethics Committee of Research Institute of Social Development, Southwestern University of Finance and Economics.

### Measures

#### Dependent variable: life satisfaction

Life satisfaction was measured following an approach to that of Cui [90], who developed 20 items based on Diener’s concept of life evaluation (past, present, and future) to measure the life satisfaction of older people. Six questions were used to measure past life satisfaction, for example, “There were few regrets in my life”; “Compared with my peers, I had done a lot of stupid things”; “Looking back on my life, I had done almost nothing right”; and “My life was very hard and lonely, I do not want to think about it”. Eight questions were used to measure present life satisfaction, for example, “Recently, I feel very happy in my life”; “I am happy with my lifestyle”; “Every day, life is very boring”; “Recently, I am prone to get angry”; “Recently, there are more sad things than happy things”. Six questions were used to measure future life satisfaction, such as “I am full of hope for the future”; “In the future, there will be many interesting and happy things around me”; “In the future, there is almost nothing that I will be able to do”; and “In the future, I will no longer be able to do anything of value”. Statements were responded to on a 5-point frequency scale ranging from 1 to 5, with 1 denoting never, 2 denoting seldom, 3 denoting sometimes, 4 denoting regularly, to 5 denoting very frequently. Responses were partially reverse-coded, with higher score indicating greater life satisfaction. The reliability factor Cronbach’s alpha was 0.821.

#### Independent variable: social trust

Social trust was measured following an approach similar to that of Lin [91]. Social trust includes interpersonal trust and institutional trust. Five questions were used to measure interpersonal trust, such as, “Do you trust your family, relatives, friends, neighbors, and strangers?” Nine questions were used to measure institutional trust, such as, “Do you trust the central government institutions, local government institutions, national people’s congress, educational institutions, medical institutions, judicial branches, Public Security Bureau, news media organization and neighborhood committee?” Responses were recorded on a five-point Likert scale, that was divided into five levels: “completely distrust,” “distrust,” “neither trust nor distrust,” “trust,” and “completely trust.” The scores were then averaged. A higher score indicates higher social trust. The reliability factor Cronbach’s alpha was 0.814.

#### Mediating variable: social network

Social network was measured following an approach to that of Lin [91]. Social network includes informal and formal social networks. The informal social network was measured on the basis of six questions, for instance, “How many children would like to give you money or gifts?” “How many relatives or friends would like to give you money or gifts?” “How many times do you contact your children in a month (meeting or by phone)?” “How many times do you contact your relatives or friends in a month (meeting or by phone)?” “How many friends or neighbors might be able to help you in a time of trouble?” and “How many neighbors are willing to treat you to a meal or do you a small favor (pick up your package, lend you a chair or supplies, etc.)?” Two questions were used to measure the formal social network, namely, “How many group activities have you participated in (community meetings, interest activities, etc.)?” and, “How many times do you meet members of the group in a month?” The scores were then averaged. A higher score indicates a wider social network. The reliability factor Cronbach’s alpha was 0.538.

#### Control variable*s:* gender, age, educational attainment, and marital status

In this study, we controlled for the following variables: gender, age, educational attainment, and marital status. We collected data of the demographic variables of gender (0 = male; 1 = female), educational attainment (0 = no formal education; 1= primary school; 2 = middle school; 3= high school; 4= vocational school; 5= college education), and marital status (0= married; 1= separated or divorced; 3= widowed). Age was a continuous variable.

### Statistical analysis

SPSS 22.0 and Amos 22.0 were used for statistical analysis. First, we used SPSS 22.0 for descriptive and correlational analyses. Second, we used Amos 22.0 for Structural equation modeling (SEM) analysis. SEM represents a comprehensive multiple regression method for explaining the relationship between observed and latent variables. SEM is a combination of measurement models, including confirmatory factor analysis models and structural models [92]. Moreover, Amos 22.0 was used to test the fitness of the entire model and the relationship between the variables. Moreover, to examine the relationship between social trust, social network, and life satisfaction, a path analysis was performed, including χ^2^ verification, and four fitness indices, namely TLI, CFI, RMSEA, and SRMR, were used to evaluate the model. Finally, the bootstrap method was used to confirm the mediating effects of social networks. The bootstrap confidence interval (CI) was set to 95%, if the CI of the indirect effect did not include zero, the mediation effect was proved to be significant.

## Results

### Descriptive results

Table 1 presents the descriptive statistics of the socio-demographic characteristics. The respondents’ average age was 72.535 years. Among 297 participants, 133 (44.1%) were men and 166 (55.9%) were women. In terms of educational attainment, 106 (35.7%) respondents had completed primary school and below, 111(37.4%), had middle and high school education and 80 (26.9%) had vocational education or above. Regarding marital status, 293 respondents were married (98.7%), 3 were separated or divorced respondents 1.0%, and 1 was widowed 0.3%.

**Table 1.** Characteristics of the study sample (n=297)

| Variables | N (%) | M (SD) |
| --- | --- | --- |
| Gender |  |  |
| Male | 133 (44.1) |  |
| Female | 166 (55.9) |  |
| Age |  | 72.535 (52.750) |
| Educational attainment |  |  |
| No formal education | 18 (6.1) |  |
| Primary school | 88 (29.6) |  |
| Middle school | 71 (23.9) |  |
| High school | 40 (13.5) |  |
| Vocational school | 57 (19.2) |  |
| College education | 23 (7.7) |  |
| Marital status |  |  |
| Married | 293 (98.7) |  |
| Separated or divorced | 3 (1.0) |  |
| Widowed | 1 (0.3) |  |

### Correlation among social trust, social network, and life satisfaction

The results of correlation analysis between the confirmed variables are shown in Table 2. The results were statistically significant (*p* < 0.01). First, social trust was significantly positively correlated with social network (*r* = 0.173). Second, social trust was significantly positively correlated with life satisfaction (*r* = 0.285). Third, social network was significantly positively correlated with life satisfaction (*r* = 0.281).

**Table 2.** Correlations between social trust, social network, and life satisfaction of older people.

| Variables | Social trust | Social network | Life satisfaction |
| --- | --- | --- | --- |
| Social trust | 1 |  |  |
| Social network | 0.173*** | 1 |  |
| Life satisfaction | 0.285*** | 0.281* | 1 |

\* $p < 0.05$ , \*\* $p < 0.01$ , \*\*\* $p < 0.001$ .

### Descriptive statistics of major variables, and reliability and validity tests

Major variables were life satisfaction, social trust, and social network. All variables were composed of latent variables. Before the SEM analysis, the standard error, Z value, *p* value, reliability, and validity of each variable were examined. All results of the regression analyses were statistically significant (*p* < 0.01). The CR (composite reliability) value is a combination of all measured variables and indicates dimensional reliability of the dimensional metrics. The higher the CR value, the higher the internal reliability. A combined reliability of 0.7 was the threshold for acceptable reliability [93] Average variance extraction (AVE) is the average ratio of the explanatory power of the latent variable to that of the measured variable. A higher AVE value represents higher convergence validity of dimensions. The AVE value should be > 0.5, and 0.36–0.5 was the threshold for acceptable validity [94]. According to Table 3, a CR value of > 0.7, and an AVE value of > 0.48 were observed. The CR and AVE values were within the acceptable numerical range.

**Table 3.**
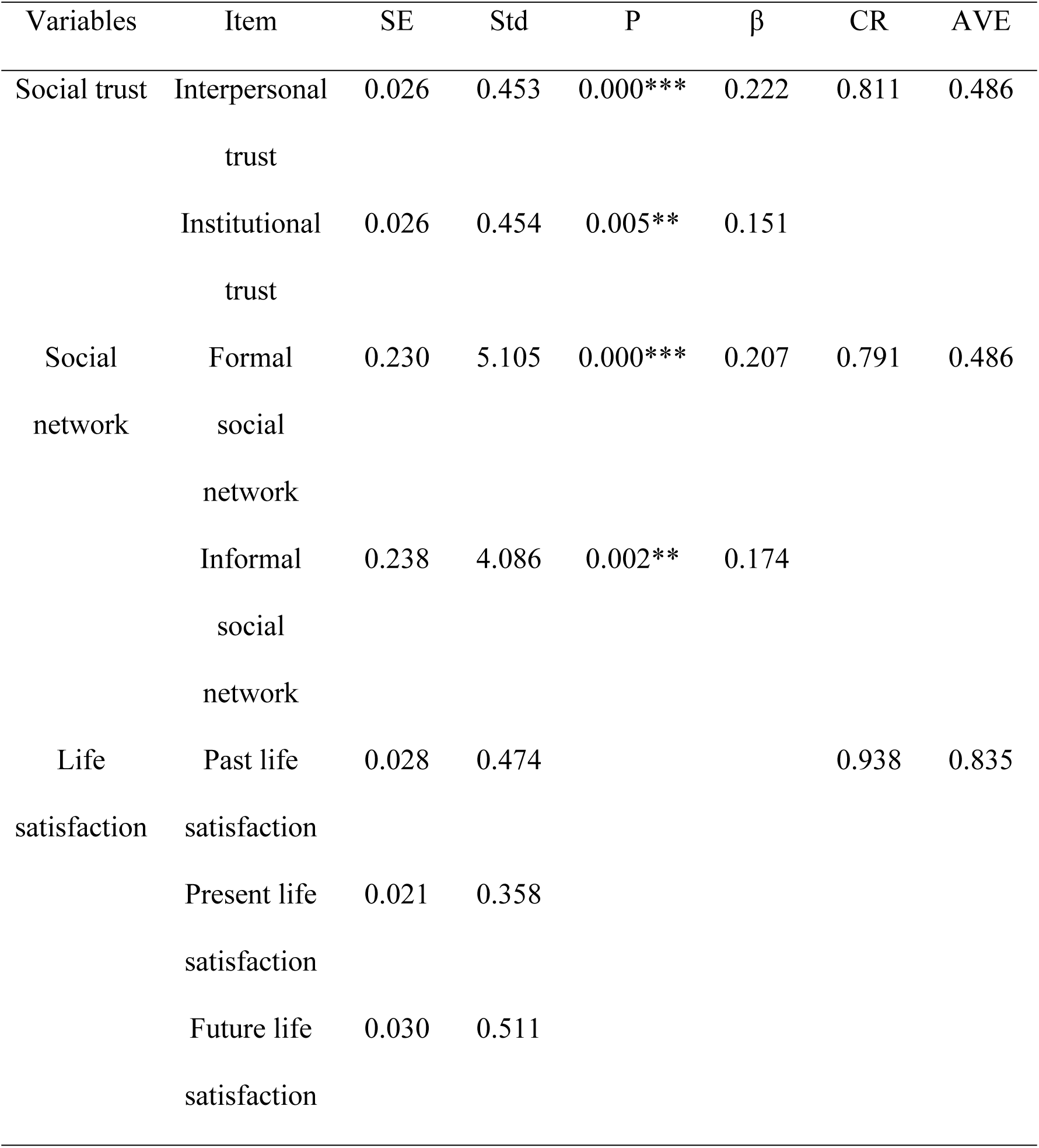
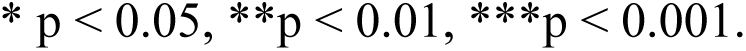
Descriptive statistics of key variables, reliability and validity test of measurement items.

### Confirmatory factor analysis model results

#### The fitness of the model

Table 4 shows the results of the test of fitness of the model for the three latent variables, and the χ^2^ value was 24.721 (*df* = 17; *p* = 0.000). TLI, CFI, and SRMR values were used to evaluate fitness. TLI value of = 0.922, and CFI value of = 0.977, that is, both values ≥ 0.90, showed a suitable fit. RMSEA value = 0.065, and RMR value = 0.0375, which were considered acceptable.

**Table 4.** The fitness of the model.

| $\chi^2$ | df | TLI | CFI | SRMR |
| --- | --- | --- | --- | --- |
| 24.721 | 17 | 0.922 | 0.977 | 0.0375 |

#### Structural model results

##### Research model regression and path analysis results

According to Table 5 and Figure. 1, the regression and all path coefficients were statistically significant (*p* <0.01). Specifically, social trust was positively related to social network. The stronger the social trust, the more beneficial it is to expand the social network (*B* =0.472; *p* <0.01). In addition, social network was positively related to life satisfaction. The more extensive the social network, the higher is the life satisfaction (*B* =0.433; *p* <0.01). Moreover, social trust positively affected life satisfaction. The stronger is the social trust, the higher is the life satisfaction (*B* =0.410; *p* <0.01).

**Fig. 1.**
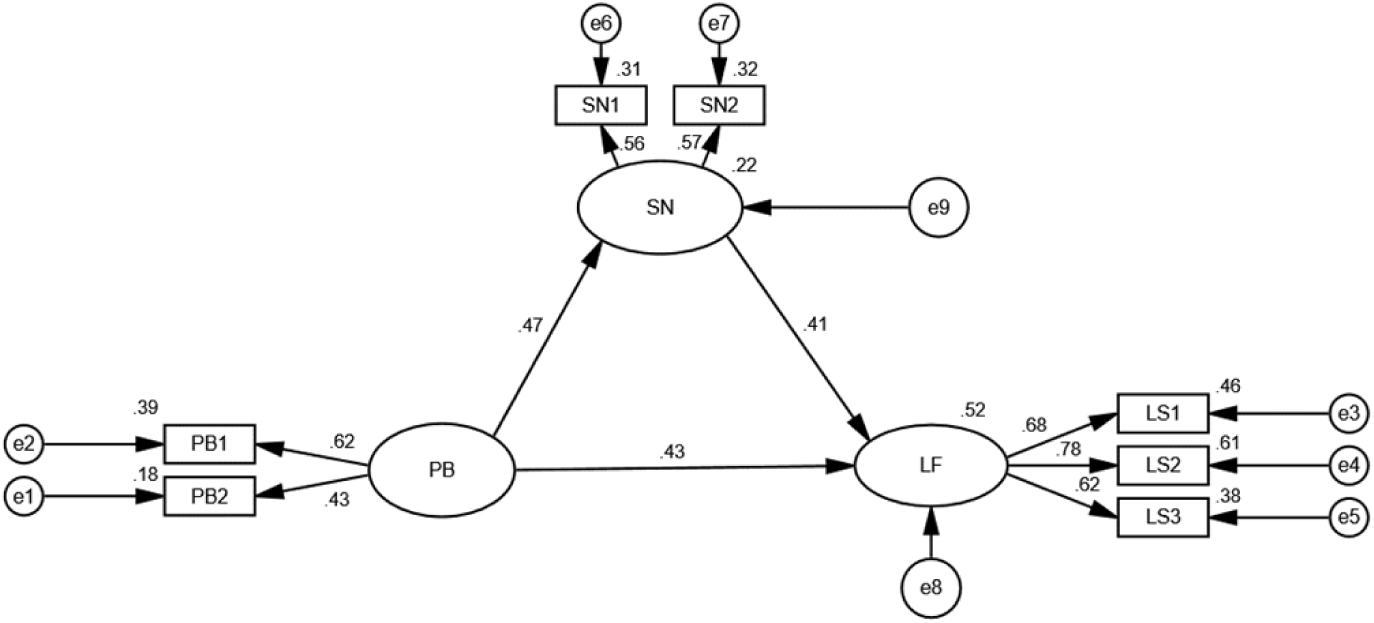
Research model. PB= social trust, SN= social network, LF= life satisfaction.

**Table 5.** Research model path analysis.

| Parameter | Estimate | SE | CR | P | Std estimate |
| --- | --- | --- | --- | --- | --- |
| PB→LF | 0.709 | 0.255 | 2.785 | 0.005** | 0.427 |
| SN→LF | 0.047 | 0.017 | 2.824 | 0.005** | 0.411 |
| PB →SN | 6.889 | 2.331 | 2.956 | 0.003** | 0.470 |

Abbreviations: PB=social trust, SN=social network, LF=life satisfaction.

### Mediating effect

Bootstrapping, was performed to verify the mediating effect. The mediation analysis found that the relationship between social trust and life satisfaction was partially mediated by social network in older people (Table 6). The total influence of positive social trust on life satisfaction was 1.030. The direct effect of social trust on life satisfaction remained significant in older people (the direct effect = 0.709), and the indirect effects of social trust on life satisfaction through social network were also significant (indirect effects = 0.321; bootstrap 95% CI = [0.051, 0.968]).

**Table 6.** Summary of mediating results.

| PB → SN → LF |  | Bootstrapping<br>bias corrected<br>95% CI |  |  |  |
| --- | --- | --- | --- | --- | --- |
| Variable<br>relationship | Effect | Lower | Upper | SE | P |
| Direct<br>effect | 0.709 | 0.115 | 1.561 | 0.325 | 0.000*** |
| Indirect<br>effect | 0.321 | 0.051 | 0.968 | 0.212 |  |
| Total effect | 1.030 | 0.612 | 1.766 | 0.286 | 0.000*** |

## Discussion

### Social trust and life satisfaction

Our study used self-evaluation to assess life satisfaction among older people. The results suggest that older people with stronger social trust have higher life satisfaction and more extensive social network. Yet our findings are consistent with previous observations, which showed that high levels of interpersonal trust and institutional trust not only reflect good-quality social relationships, but also promote meaning in life. These in turn affect one’s subjective life satisfaction evaluation [95, 88]. A researcher used Statistics Canada’s 2003 General Social Survey data, which were obtained by interviewing 24,951 people aged 15-80 years. The results revealed that both institutional trust and interpersonal trust are related to life satisfaction [12].

### Social network and life satisfaction

As hypothesized, we also found that social network was positively related to life satisfaction in older people. This is similar to the results of previous studies, which showed that social resources embedded in social networks play a crucial role in the life of older people living in rural areas of China [96, 97]. In China, social interaction has developed over thousands of years, staring with an agricultural civilization rooted in the small-scale, local peasant economy. It emphasizes human relationships among family members, relatives, and neighbors [16]. In particular, after retirement, older people face a loss of major organizational networks and rely more on individual social networks [63]. Among U.S. residents aged 65 years and older, friendship-centered social networks were related to better well-being than family-centered social networks [52]. The difference is a researcher highlighted the significance importance of family support and cognitive community social capital in improving the life satisfaction of Chinese urban older adults [98]. With age, older people become more dependent on social relationships to maintain their well-being, and the trust is the basis and prerequisite for building these social relationships [99]. Society is networked, and in China, those well-connected are happier than those who are not [50]. In summary, good interpersonal relationships led to attainment of life satisfaction [100–102]. This is, because, social relationships can enhance social identity, thereby providing instrumental and affective supports for members of social groups who share resources. This could be explained in the context of the socioemotional selectivity theory [62], that is, older people are likely to choose social networks with strong social trust and interactive relationships. They regard the social network as a means to improve life satisfaction [30]. It is the perception of the quality of social networks rather than the perception of the quantity of social networks that is relevant to reporting better well-being [103, 104]. Moreover, in China, older people are expected to play valuable social roles, and social networks offer opportunities to the elderly people to realize their self-worth, which are conducive to maintaining good interpersonal relationships, thus increasing the sense of happiness [17, 31].

### Social trust and social network

We also found that social trust was positively correlated with the social network in older people. This result is similar to those of previous studies. Social ties and interactions are strengthened by the accumulation of social trust [73]. Frequent social interactions are conducive to the expansion of social networks [105]. Good social relationships are crucial for older people as they constantly require support in the form of positive social interactions. These interactions also act as a means to satisfy their instrumental and affective needs, such as obtaining help, and the feeling of being needed and appreciated [106, 82]. A stronger social trust, is more conducive to the strengthening of social ties and improving social participation [107]. If social participation of older people is higher, they receive more opportunities to strengthen their social interactions because they may have more chances to interact face-to-face with others. All of these interactions could contribute to the development of older people’s social networks [108,109].

### Mediation effect of social network on the relations between social trust and life satisfaction

As hypothesized, this study revealed that social network mediates the effect of social trust on life satisfaction in older people. This finding is in agreement with that of Kawachi’s study [71]. Accordingly, higher social trust is more conducive to the construction of the social network, and more social support can be gained and requirements are met in a better way if the social network is broad, which in turn can increase, life satisfaction of older people. Moreover, this result could be explained by social capital theory. Although many researchers have defined social capital, this paper adopts Lin’s and Adler and Kwon’s definition of social capital: “Social capital is an aggregation in a specific network composed of mutual understanding and mutual trust, and through social interaction, resources embedded in a social network are captured [110, 111].” Different from other capital, social capital exists in social relations that promote the formation of social trust and networks, thus improving the life satisfaction of the elderly people [112, 113]. Social trust enhances people’s interaction and expands their social network, which is the basis for the premise of interpersonal communication of elderly people. Regarding why social trust is related to health, researchers have reached consensus that trust is a key indicator of social ties and social network [114]. In turn, social network benefits health through obtainment of social resources and social support, which are closely associated with individuals’ health behaviors and well-being [97, 115]. Webster et al. (2015) predicted that social trust could affect the health of the elderly people through social networks [116]. In addition to the mediating roles of social networks in social trust and life satisfaction, political interest plays mediating role in social trust and well-being in older adults [117]. Another review of studies with older adults suggested that more recreational participation and social trust were associated with health in China, and good health predicts high life satisfaction [118, 119].

Additionally, social trust belongs to the category of cognitive social capital. By contrast, social network belongs to the category of structural social capital [120]. At the level of structural social capital, a strong link exists between social capital and happiness. Because structural social capital relies on social interactions, older people are encouraged to form social networks and, over time, to strengthen social relationships, which could enhance the sense of belonging and social inclusion, and thus improve their quality of life [121]. This study has demonstrated that social network plays a mediating role in the relationship between social trust and life satisfaction. In other words, structural social capital mediates the effect of cognitive social capital on life satisfaction in older people in China. The relationships between cognitive capital, structural capital and life satisfaction have been previously documented [23, 122]. Kim et al (2020) reported that social participation and both cognitive social capital (feelings of trust) and structural social capital (neighborhood connections) could predict one’s happiness [123]. However, different from the conclusion of this paper, using a sample of Chinese urban older adults, Sun and Lu (2020) confirmed that cognitive social capital had a mediation effect on the association between structural social capital and life satisfaction [95].

## Strengths and limitations

Our study had several strengths. First, to the best of our knowledge, this study is the first to reveal the mediating effect of social network on the relationship between social trust and life satisfaction of older people in China. Our findings provide a new perspective for enhance the life satisfaction of older people. Second, this study used social capital theory to explain the relationship among social trust, social network, and life satisfaction. We found that structural social capital mediates the relationship between cognitive social capital and life satisfaction. Our findings extended the practical application of social capital theory. The findings proved new empirical evidence to the social capital theory from the Chinese perspective. This paper sheds some light on this topic with reference to Chinese older people, thereby enriching the empirical research on social trust, social network, and life satisfaction. However, the study some limitations. First, the data were cross-sectional, exploring causal relationships between variables was difficult. Therefore, a future study could collect long-term data and examine longitudinal changes and causality. Second, data in this study were limited to the surveyed older people residing in Chengdu, Sichuan Province. Given the vast size of China, and the unique demographic characteristics of each region, the differences between rural and urban areas, and between different provinces need to be investigated.

## Conclusion

This study examined the associations between social trust and life satisfaction and the mediation effect of social network on these associations among older Chinese people. Our study findings confirmed all the hypotheses made in the paper. We found that social trust is the premise and basis for building social network. Social network is a carrier for older people to interact and communicate with others. Older people with strong social trust can obtain more instrumental and emotional support to improve their life satisfaction through different social networks. In conclusion, this study exploring the mediating effect of social network provides knowledge essential for ensuring a happy old age. The study results can help advance psychological research on life satisfaction of older people and suggest new intervention strategies in this field.

## Abbreviations

PB: Social trust
SN: Social network
LF: life satisfaction

## Acknowledgments

The authors would like to express their gratitude to all the older people who participated in this study.

## Author’s contributions

HXY conceived and designed the study, collected and analyzed the data, and wrote the initial draft. LHM and YZY designed the research framework, methodology, data analysis and writing-review. LSH and XCX helped to obtain funding. LSH were involved in the research for the study and revised the manuscript. All authors have read and approved the final manuscript.

## Availability of data and materials

Data are available from the authors. Any interests, please contact the author of correspondence.

## Declarations

### Ethics approval and consent to participate

Ethical approval for this study was obtained from the Humanities and Law School, Chengdu University of Technology (No.20230116). All older participants received written informed consent before participating in this study, anonymity was maintained throughout the study and all data collected was securely stored to protect the privacy of participants. In addition, all the methods used in our research were implemented in accordance with the guidelines for Human research of the Helsinki Declaration.

### Consent for publication

Not applicable

## Footnotes

1 Besides the definition stated above, an alternative definition of an ageing society is one with the population above the age of 65 accounted for 7% of the total population. This particular portion of population in China reached 13.5% in 2020 (The Seventh Census, National Bureau of Statistics of China)

